# Metabolic signature and proteasome activity controls synovial migration of *CDC42^hi^*CD14^+^ cells in rheumatoid arthritis

**DOI:** 10.1101/2023.06.15.23291416

**Authors:** Eric Malmhäll-Bah, Karin M.E. Andersson, Malin C. Erlandsson, Sofia T. Silfverswärd, Rille Pullerits, Maria I. Bokarewa

## Abstract

**Objective:** Activation of Rho-GTPases in macrophages causes inflammation and severe arthritis in mice. In this study, we explore if Rho-GTPases define the joint destination of pathogenic leukocytes in rheumatoid arthritis (RA) and how JAK inhibition mitigates these effects.

**Methods:** CD14^+^ cells of 136 RA patients were characterized by RNA-sequencing, and cytokine measurement to identify biological processes and transcriptional regulators specific for *CDC42*^hi^CD14^+^ cells, which were summarized in a metabolic signature. Effect of hypoxia, and IFN-γ signaling on the metabolic signature of CD14^+^ cells was assessed experimentally. To investigate its connection with joint inflammation, the signature was translated into the single cell characteristics of *CDC42*^hi^ synovial tissue macrophages. Sensitivity of the metabolic signature to the RA disease activity and treatment effect was assessed experimentally and clinically.

**Results:** *CDC42*^hi^CD14^+^ cells carried the metabolic signature of genes functional in the oxidative phosphorylation and proteasome-dependent cell remodeling, which correlated with the cytokine-rich migratory phenotype and antigen presenting capacity of these cells. Integration of *CDC42*^hi^ CD14^+^ and synovial macrophages marked with the metabolic signature revealed the important role of the interferon-rich environment and immunoproteasome expression in homeostasis of these pathogenic macrophages. The *CDC42*^hi^CD14^+^ cells were targeted by JAK-inhibitors and responded with downregulation of immunoproteasome and MHC-II molecules, which disintegrated the immunological synapse, reduced cytokine production and alleviated arthritis.

**Conclusion:** This study shows that the CDC42-related metabolic signature identifies the antigen-presenting CD14^+^ cells that migrate to joints to coordinate autoimmunity. Accumulation of *CDC42*^hi^CD14^+^ cells disclose patients perceptive to JAKi treatment.

## Introduction

Monocytes are the crucial innate effectors in pathogenesis of the canonical inflammatory joint disease rheumatoid arthritis (RA). Monocytes are heterogenous population that in healthy maintain immune homeostasis of the joint tissues supporting renewal and anti-microbial protection^1,2^. Migration and infiltration of pro-inflammatory monocytes into synovial tissue is an early sign of RA^3-5^. Pro-inflammatory monocytes differentiate to macrophages and play the key role in the propagation of synovial inflammation by maintaining a continuous influx of leukocytes into the joint compartment of RA patients^3^.

Monocytes together with fibroblast-like synoviocytes, present the main pool of antigen presenting cells in the joint cavity^6,7^. During inflammation, the number of CD14^+^ monocytes increase in circulation of RA patients and shows higher oxygen consumption rate and number of mitochondria^8^. In contrast to healthy, the inflamed synovium is hypoxic^9,10^, which transforms the monocyte subsets with respect to the surface receptor phenotype, antigen presenting capacity and cytokine production.

The key in the antigen presentation process is the Generation of peptides for loading onto the major histocompatibility complex (MHC) is the key in the antigen presentation to CD4^+^ and CD8^+^ T cells. Although fragmentation of proteins in the proteasome complex followed by loading within the endoplasmic reticulum has initially been identified in MHC class I receptors (MHC-I) interacting with CD8^+ 11^, this process is equally relevant for MHC class II receptors (MHC-II) and CD4^+^ cells^12^. The proteasome complex is an integral part of cell homeostasis. The constitutive 26S proteasome consists of a barrel-shaped catalytic core and a regulatory lid associated to it. To be disintegrated by constitutive proteasome, the protein needs to be ubiquitinated, unfolded by the regulatory lid, and catalyzed into peptides. All these events are dependent on the energy released by hydrolysis of ATP^13,14^. To generate peptides suitable for the antigen presentation by MHC complexes, the proteasome exchanges its composition. Specifically, the regulatory lid is replaced by an open-gate proteasome activator complex PA28, and the catalytic core replaces three of its catalytic β-subunits with endopeptidases encoded by *PSMB8, PSMB9*, and *PSMB10*. These changes recognize the immunoproteasome complex, which has an ATP-independent dynamics of protein degradation and results in generation of more hydrophobic peptides^15^. Several conditions induce a transition of the constitutive to immunoproteasome, among those are exposure to pro-inflammatory cytokines interferons (IFN-α,β,γ) and TNF-α, and stress factors including oxygen deprivation, exposure to toxins, etc.^16^.

In our previous studies, we found that conditional knockout of GGTase-I protein (GLC) in mouse macrophages caused a symmetrical arthritis and skeletal damage in small joints, which was morphologically identical to human RA. The GLC macrophages were recognized by an accumulation of the activated forms of RAC1, RHOA and CDC42 GTPases^17,18^. Rho-GTPases are signal transducers that regulate the actin cytoskeleton and cell migration, but have recently been implicated in vital cellular functions of metabolic regulation, differentiation, and cell cycle control^19,20^. Additionally, Rho-GTPases are engaged in the process of antigen presentation. For example, it has been shown that constitutively active CDC42 mediates antigen presentation between dendritic cell and T cells^21^, while *CDC42* deficiency led to a reduction in lysosomal content and upregulation of dysfunctional invariant chain^22^. Concordantly, we demonstrated that GLC macrophages efficiently transduced upregulation of Rho-GTPases to CD4^+^T cells and created IFN-γ rich environment. This resulted in an excessive egress of the regulatory T cells to periphery that later acquired invasive phenotype and migrated into joints causing arthritis^23^. Similar to the *GLC* macrophages, deletion of *Pggt1b* in CD4^+^ cells resulted in hyperactivated Rho-GTPases and aggravated autoimmune colitis^24^. Modulation of Rho-GTPases in experimental autoimmunity efficiently changed disease severity. Deletion of the activated Rac1 and RhoA Rho-GTPases in GLC macrophages resulted in alleviation of arthritis in mice^23,25^. Consistently, RhoA-deficiency in CD4^+^ cells alleviated autoimmune encephalomyelitis^26^, while the gain-of-function mutation in the *RHOA* gene induced a T cell dependent autoimmunity^27^. The conditional deletion of CDC42 demonstrated that this Rho-GTPase was the main mediator of interaction between macrophages and CD4^+^ cells, which promoted T cell expansion and invasion^23,28,29^. This exemplifies importance of Rho-GTPases in regulation of the experimental autoimmunity, while information about the role of Rho-GTPases in human disease remains sparse.

In this study, we investigated the metabolic signature of *CDC42*^*hi*^CD14^+^ cells and its connection with synovial macrophages of RA patients. Integrating independent sample collections and single-cell resolution, we explored the process of *CDC42*^*hi*^CD14^+^ cell transformation in synovium with focus on antigen presentation and proteasome-dependent protein remodeling and demonstrated these mechanisms to be the key to pathogenic synovial tissue macrophages in RA. Finally, we examined the ability of anti-rheumatic treatment affecting these mechanisms and exposed their revertible nature.

## Material & Methods

### Patients

Blood samples of 59 females RA patients were collected at the Rheumatology Clinic, Sahlgrenska Hospital, Gothenburg. Clinical characteristics of the patients are shown in Tab. S1. All RA patients fulfilled the EULAR/ACR classification criteria^30^ and gave written informed consent before the blood sampling. The study was approved by the Swedish Ethical Review Authority (659-2011) and done in accordance with the Declaration of Helsinki. The trial is registered at ClinicalTrials.gov (ID NCT03449589). In this study, CD14+ and CD4+ cells from RA patients were used for RNA-seq, qPCR, ELISA, and culturing experiments in hypoxic environment with and without IFN-γ stimulation as well as culturing experiments with JAK-inhibitors. Additional RNA-seq dataset of CD14^+^ cells from 77 RA patients with active disease and naïve to treatment was accessed (accession no. GSE138747)^31^ from the Gene Expression Omnibus database (GEO, RRID:SCR_005012). The corresponding clinical data of these 77 patients was kindly provided by Dr. Tao and is shown in Tab S1. Lastly, scRNA-seq dataset of synovial tissue HLA-DR^+^CD11b^+^ macrophages sorted by flow cytometry from the arthroscopic biopsies of 25 RA patients^32^ was accessed from the EMBL’s European Bioinformatic Institute depository (accession no. E-MTAB-8322, RRID:SCR_004727).

### Isolation of CD14^+^ and CD4^+^ cells

Human peripheral blood mononuclear cells were isolated from venous peripheral blood by density gradient separation on Lymphoprep (Axis-Shield PoC As, Dundee, Scotland). CD4+ cells were isolated by positive selection (11331D; Invitrogen, Waltham, Massachusetts, USA), and cultured (1.25x106 cells/ml) in wells coated with anti-CD3 antibody (0.5 mg/ml; OKT3, Sigma-Aldrich, Saint Luis, Missouri, USA, RRID: AB_2619696), in RPMI medium supplemented with 50μM b2-mercaptoethanol (Gibco, Waltham, Massachusetts, USA), Glutamax 2mM (Gibco), Gentamicin 50μg/ml (Sanofi-Aventis, Paris, France) and 10% fetal bovine serum (Sigma-Aldrich) at 37°C in a humidified 5% CO2 atmosphere for 48 hours. CD14^+^ were subsequently purified from the remaining cell mixture by positive selection (17858; Stem Cell Technologies, Vancouver, Canada) and cultured in the same medium and conditions as the CD4^+^ cells but stimulated with LPS (5 μg/ml; Sigma-Aldrich). Culture supernatants were collected for analysis of cell products by ELISA.

### Culturing experiments CD14^+^ cells

Human peripheral blood mononuclear cells were isolated from venous peripheral blood by density gradient separation on Lymphoprep (Axis-Shield PoC As). CD14^+^ cells were isolated by positive selection (Stem Cell Technologies), and cultured (1.25x106 cells/ml) in RPMI medium supplemented with 50μM b2-mercaptoethanol (Gibco), Glutamax 2mM (Gibco), Gentamicin 50μg/ml (Sanofi-Aventis) and 10% fetal bovine serum (Sigma-Aldrich). In the experiment determining effect of hypoxia and IFN-γ cell stimulation was performed by the addition of LPS (5 μg/ml; Sigma-Aldrich) and IFN-⍰(50 ng/ml; Peprotech, Cranbury, NJ, USA) in either normoxic (95% air, 5% CO2) or hypoxic (1% O2, 5% CO2, and 94% N2) conditions over 48 hours at 37°C. To determine the effect of JAK-inhibitors cells were stimulated by the addition of LPS (5 μg/ml; Sigma-Aldrich) with or without tofacitinib (10 μM; CP-690550; Selleck Chemicals, Houston, Texas, USA).

### Transcriptional sequencing (RNA-seq)

RNA was prepared with the Norgen Total RNA purification kit (37500; Norgen Biotek, Ontario, Canada). Quality control was done with a Bioanalyzer RNA6000 Pico on an Agilent2100 (Agilent, St.Clara, CA, USA, RRID:SCR_019715). Deep sequencing was done by RNA-seq (Hiseq2000; Illumina, San Diego, Kalifornien, USA, RRID:SCR_020132) at the LifeScience Laboratory, Huddinge, Sweden. Raw sequence data were obtained in Bcl files and converted to fastq text format with bcl2fastq. RNA-seq results were validated by qRT-PCR as described below.

### Conventional qPCR

RNA was isolated with the Total RNA Purification Kit (37500; Norgen Biotek). RNA concentration and quality were evaluated with a NanoDrop spectrophotometer (Thermo Fisher Scientific, RRID:SCR_018042) and Experion electrophoresis system (Bio-Rad Laboratories, RRID:SCR_019691). cDNA was synthesized from RNA (200 ng) with the High-Capacity cDNA Reverse Transcription Kit (4368814; Applied Biosystems, Foster City, CA, USA). Real-time amplification was done with RT2 SYBR Green qPCR Mastermix (330522; Qiagen, Hilden, Germany) and a ViiA 7 Real-Time PCR System (Applied Biosystems, RRID:SCR_023358) as described^33^. Melting curves for each PCR were performed between 60 and 95 °C to ensure specificity of the amplified product. All samples were run in duplicate with ACTB (beta-actin) as a reference gene and with a negative control. Expression levels of target genes were normalised to ACTB to obtain the difference in cycle threshold (dCt) using the QuantStudioTM Real-time PCR software (v1.3; Applied Biosystems). The relative quantity (RQ) was calculated using the ddCt method. Primers used are shown in Supplementary Table S2.

### ELISA Cytokine measurement

Cytokine levels were measured with a sandwich enzyme-linked immune assay (ELISA) as below. Briefly, high-performance 384-well plates (Corning Plasticware, Corning, NY, USA) were coated with capture antibody, blocked, and developed according to the manufacturers’ instructions. Developed plates were read in a SpectraMax340 Microplate reader (Molecular Devices, San Jose, CA, USA, RRID:SCR_020303) at the dual wavelength of 450/650 nm, and absolute protein levels were calculated after serial dilutions of the recombinant protein provided by the manufacturer. The following reagents were used, for IFN-(detection limit 3 pg/ml, PelikineM1933, Sanquin, Amsterdam, The Netherlands, RRID:AB_2935684), TNF-α (detection limit 15.6 pg/ml, DY210, R&D Systems, RRID:AB_2848160), IL-1β (detection limit 3.9 pg/ml, DY201, R&D Systems, RRID:AB_2848158), IL-6 (detection limit 9.4 pg/ml, DY206, R&D Systems, RRID:AB_2814717) CXCL8 (detection limit 31.2 pg/ml, DY208, R&D Systems), and IL-10 (detection limit 15 pg/ml, DY217B, R&D Systems, RRID:AB_2927688).

### RNA-seq analysis

Transcripts were mapped with the UCSC Genome Browser using the annotation set for the hg38 human genome assembly and analyzed with the core Bioconductor packages in RStudio (v 4.1.1, RRID:SCR_000432). DEGs were identified with DESeq2 (v 1.26.0, RRID:SCR_015687)^34^. Genes were considered differentially if p_nominal_<0.05. ComplexHeatmap (v 2.8.0, RRID:SCR_017270) was used to cluster genes and construct heatmaps. Pathway enrichment analysis was performed using g:Profiler web client (ELIXIR Infrastructure, RRID:SCR_006809) ^35^, with a term size filter of 1000. Enrichment of transcription factor targets were determined using the GSEA web client (Broad Institute, RRID:SCR_016863)^36^, testing genes against the TFT collection (GTRD^37^ and LEGACY).

### Single-cell RNA-sequencing analysis

Synovial tissue macrophages of interest (STM) were identified in scRNA-seq dataset obtained by Alivernini, MacDonald (32). Utilizing Seurat R package (v 4.1.0, RRID:SCR_016341)^38^ cells with unique feature counts over 4,500 or less than 500 and >20% mitochondrial counts were analyzed. The dimensionality of dataset was determined with the ElbowPlot() function. Cells in dataset were clustered into 15 clusters by Unsupervised Uniform Manifold Approximation and Projection (UMAP). CDC42hiMetSighi STM cluster was identified by summation of the aggregate expression of each gene in the MetSig (*ATP5BP, COX7A2, PSMB6, PSME3, GTF3C6, and GTF2E2*), by extracting with the AggregateExpression() function and dividing by number cells of each cluster. The FindAllMarkers() function was used to determine the markers of the CDC42hiMetSighi STM cluster, using the default Wilcoxon rank sum test method with the remaining cluster as comparison. Genes were considered markers if |log2FC|>0.25 and minimum feature percentage detection at 25 %.

### Regression model

A linear equation between the point representing the maximums and the point representing the minimums of the variables of interest. Using the equation, a predicted value was calculated, and samples were considered inside the model if the absolute value of the difference between the predicted and real value was less than 1.

### Principal component analysis

Principal components analysis was performed using PCA() function of R package FactorMineR (v 2.6, RRID:SCR_014602) and visualized using the fviz_pca_biplot() function of factoextra (v 1.0.7, RRID:SCR_016692).

## Results

### Enriched oxidative phosphorylation and transcriptional regulation in *CDC42*^*hi*^CD14^+^ cells

To investigate the complete transcriptome of *CDC42*^hi^CD14^+^ cells, we utilized 135 RNA-seq datasets of blood CD14^+^ cells from two independent cohorts of RA patients. The cells with *CDC42* expression above the mean of each cohort (*CDC42*^hi^CD14^+^ cells) (Fig. S1B) were significantly enriched with other canonical Rho-GTPases *RAC1* and *RHOA* (Fig. 1B), which indicated phenotypic identity of CD14^+^ cells of the BiOCURA and NeumRA cohorts. The genes differentially expressed (DEG, normalized p-value<0.05) in *CDC42*^*hi*^CD14^+^ cells were compared. The analysis showed that 2211 DEGs were common for the BiOCURA and NeumRA cohorts and comprised 71 % and 24 % of the complete set of DEGs within each cohort, respectively (Fig. 1A). To enrich for similarity, we performed an unsupervised co-expression-based clustering of DEGs within each cohort. The pathway enrichment analysis of the common DEGs revealed that the *CDC42*^hi^CD14^+^ cells were engaged in the processes of *Leukocyte Migration* (GO:0050900, FDR=3.65e^-06^) and *Chemokine Signaling* (KEGG:04062, FDR=1.27e^-05^), which are among the best characterized functions of the Rho-GTPases^19^. Additionally, the DEG regulated energy supply through control of the *Oxidative phosphorylation* (KEGG:00190, FDR=8.18e^-05^), which supported *Actin Cytoskeleton* (GO:0015629, FDR= 4.69e^-05^) and *Proteasome complex* function (GO:0000502, FDR=4.38e^-03^), followed by *Transcription* (GO:0006366, FDR=2.91e^-07^) and *Translation* (GO:0006412, FDR= 2.04e^-14^) (Fig. 1C). This illustrated ability of Rho-GTPases to orchestrate a chain of cell functional changes.

**Figure 1.**
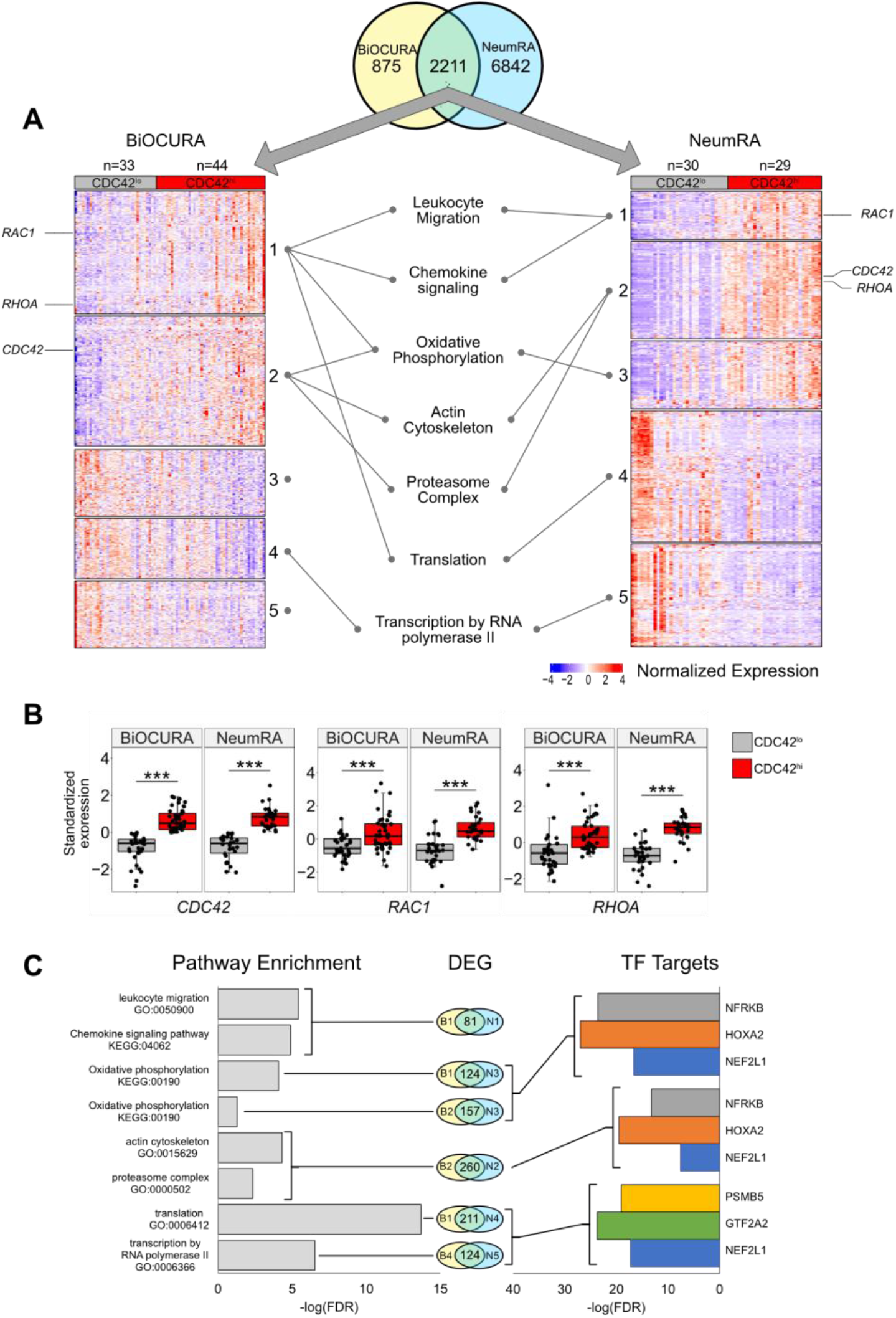
Transcriptional characteristics of CDC42^hi^CD14^+^ cells of two independent RA cohorts. Whole-genome RNA was analyzed by sequencing in blood CD14^+^ cells of RA patients (BiOCURA cohort, n=77. NeumRA cohort, n=59). Comparison between *CDC42*^*hi*^ and *CDC42*^*lo*^ (by mean expression) CD14^+^ cells was done by DESeq2 and identified differentially expressed genes (DEG, nominal p<0.05). Clustering of DEG was done by k-means. **(A)** Heatmap of DEG common for both cohorts. **(B)** Box plots of Rho-GTPase CDC42, RhoA and Rac1 expression after z-transformation. (***) indicates p-value<0.001. **(C)** Bar plots of the false discovery rate (FDR) for the GO biological processes and transcription factor (TF) targets enriched with the DEG common for each cluster.

With focus on these processes, we performed a search for the upstream transcriptional regulators of the DEGs within individual clusters using the TF target collection in the Molecular Signature Database (MSigDB, GSEA, Broad Institute). Consistent with the results of the pathways analysis, we observed an enrichment for the gene targets of the Nuclear Factor Erythroid 2-Related Factor 1 (NFE2L1, n=283), Homeobox A2 (HOXA2, n=219), and Nuclear Factor Related to Kappa-B Binding Protein (NFRKB, n=228) TF families (Fig. 1C. Fig. S1C). Together, these TFs regulated the oxidative phosphorylation (OXPHOS) through an antioxidant response motif and DNA remodeling and repair^39-41^ during the actin cytoskeleton and proteasome complex function (Fig. 1C).

Since the covariance clustering demonstrated that *CDC42*^*hi*^CD14^+^ cells were characterized by activation of OXPHOS, we performed an in-depth analysis of the individual electron transport chain (ETC) complexes and the tricarboxylic acid (TCA) cycle (Fig. 2A). We found that a substantial number of the ETC genes were significantly enriched in *CDC42*^*hi*^CD14^+^ cells of the BiOCURA and NeumRA cohorts. Furthermore, many of those DEG were direct targets of NFE2L1, HOXA2 and NFRKB and coded for components of the ATP Synthase complex (*ATP5PO, ATP5PF, ATP5PB, ATP5F1E*), NADH:ubiquinone Oxidoreductase complex (Complex I, *NDUFA4, NDUFA5, NDUFB5, NDUFB6, NDUFS4, NDUFV2*), the coenzyme Q-cytochrome c reductase complex (Complex II, *UQCRB, UQCRC2*) and the Cytochrome Oxidase subunit 7 (Complex IV, *COX7A2L, COX7B, COX7C)* (Fig. 2A,D). Additionally, the *CDC42*^*hi*^CD14^+^ cells had significantly upregulated NFRKB targets in the glucose metabolism including the glucose transporter gene *SLC2A1*, and *SUCLG1* and SUCLG2 that catalyzes the conversion of succinyl CoA to succinate (Fig. S2A). NFE2L1, NFRKB and HOXA2 control the protein remodeling including the proteasomal degradation (GO:0000502, FDR=4.83e^-03^) and the RNA polymerase synthesis (GO:0006366, FDR=2.91e^-07^) (Fig. 1C, Fig. 2B,C). The expression of proteins forming the 26S proteasome was significantly higher in *CDC42*^*hi*^ cells of both cohorts (Fig. 2E), consistent with the enhanced protein remodeling process in those cells. This included PA28-specific subunit *PSME3*, the ATPase 1 protein coded by *PSMC1*, components of the α- and β-rings of the catalytic core including *PSMB6 and PSMB7*, and components of the non-ATPase regulatory lid *PSMD7, PSMD8, PSMD10 and PSMD14*, many of which were under transcriptional control of NFE2L1^39^. Analogously, the general transcription factors (GTF) that mobilize the RNA polymerase complexes to chromatin to initiate transcription were among the transcriptional targets of NFE2L1, HOXA2 and NFRKB upregulated in *CDC42*^*hi*^CD14^+^ cells. Notably, the proteasome subunit PSMB5 and the GTF member GTF2A2 had an independent, often functionally paired, transcriptional control of translation through the genes coding for AP-1 TFs, ribosomal and histone proteins, and alpha-tubulin that enables RNA binding activity (Fig. S2B-C).

**Figure 2.**
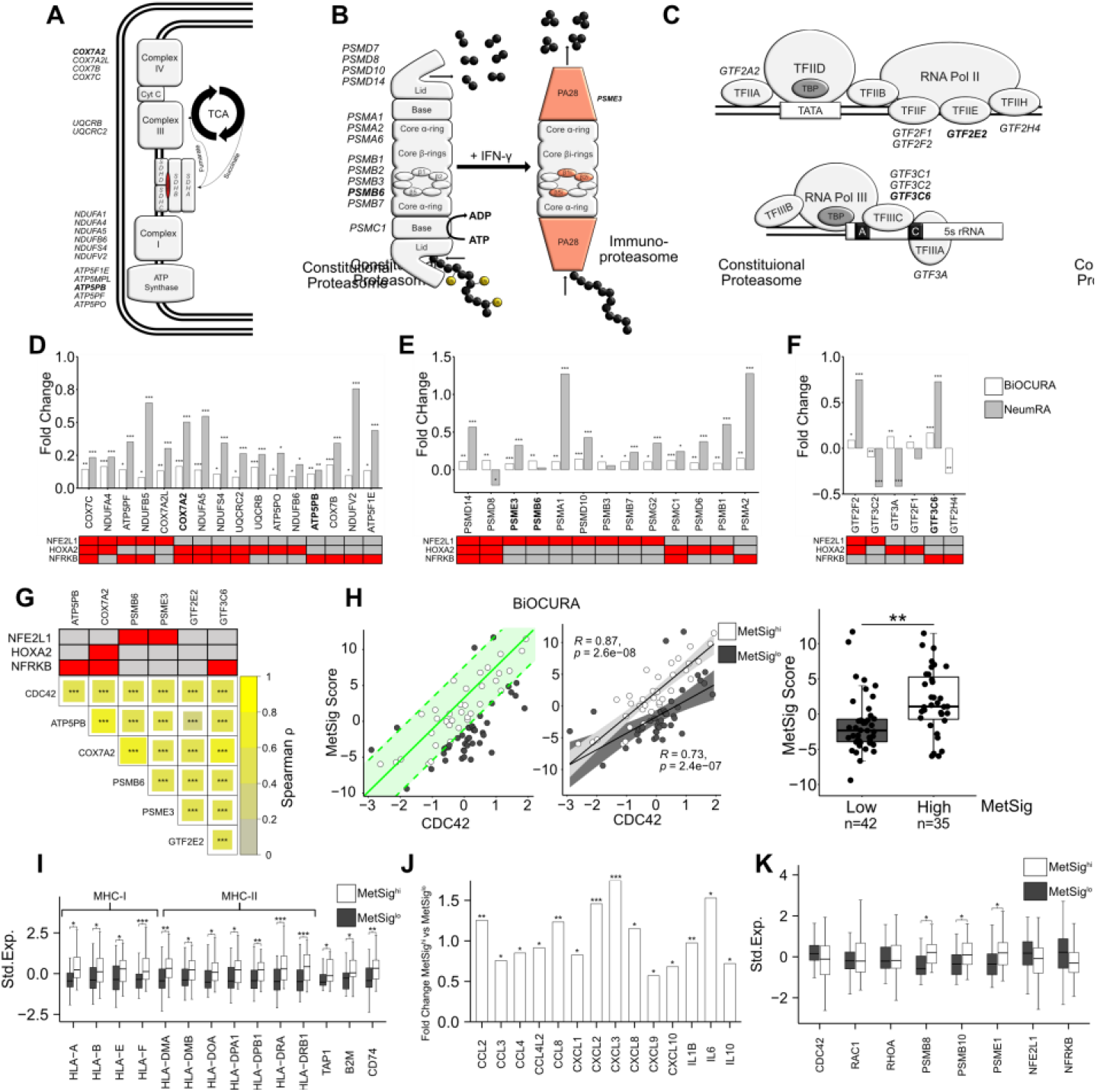
CDC42-related metabolic signature of CD14^+^ cells. **(A)** Complexes in the Electron Transport Chain (ETC) integrated in the inner mitochondrial membrane and the tricarboxylic acid cycle (TCA). **(B)** Composition of the constitutional proteasome and immunoproteasome complexes. **(C)** Composition of the Transcription Initiation complexes around Polymerase II and Polymerase III. Genes differentially expressed in *CDC42*^*hi*^CD14^+^ cells are indicated. **(D-F)** Bar plot of the gene expression difference in fold change (FC) between *CDC42*^hi^ and *CDC42*^lo^ CD14^+^ cells of BiOCURA (white bars) and NeumRA cohorts (grey bars). A net of transcriptional regulators for each gene is shown in red. **(G)** Heatmap of Spearman correlation between expression of *CDC42* and the metabolic signature (MetSig) genes. A net of transcriptional regulators for each gene is shown in red. **(H)** Scatter plot of relationship between the MetSig and *CDC42* expression. Green line indicates the linear regression model between the parameters. CD14^+^ cells within one standard deviation (green area) from the model are shown by open circles. Spearman correlation between the MetSig and *CDC42* expression within the groups. Box plot of the MetSig for CD14^+^ cells within (open circles, MetSig^hi^) and outside (solid circles, MetSig^lo^) the regression model. P-value is calculated by Wilcoxon rank sum test. (**) indicates p-value<0.01. **(I)** Box plot of genes engaged in antigen presentation in MetSig^hi^ (white bars) and MetSig^lo^ (grey bars) CD14^+^ cells of BiOCURA cohort. Gene expression is shown after z-transformation **(J)** Bar plot of the chemokine and cytokine gene expression difference in fold change (FC) between MetSig^hi^ and MetSig^lo^ CD14^+^ cells of BiOCURA cohort. **(K)** Box plot of gene expression for immunoproteasome subunits and transcription factors in MetSig^hi^ (white bars) and MetSig^lo^ (grey bars) CD14^+^ cells. P-values are calculated by DESeq2 test. (*) indicates p-value<0.05. (**) indicates p-value<0.01. (***) indicates p-value<0.001.

Taken together, the TF target analysis demonstrated that *CDC42*^*hi*^CD14^+^ cells are programmed via TFs NFE2L1, HOXA2 and NFRKB to mediate strong connection between Rho-GTPases, OXPHOS activity and proteasome-dependent protein remodeling.

### Metabolic signature defines the antigen presenting and migratory phenotype of *CDC42*^*hi*^CD14^+^ cells

Based on the knowledge gained with the analysis of the biological processes, we identified a set of internally related genes characteristic for the metabolic signature (MetSig) of *CDC42*^*hi*^CD14^+^ cells and representing OXPHOS, and proteasome-dependent remodeling. To be included in this *CDC42*^*hi*^ signature, the gene should be significantly upregulated in *CDC42*^*hi*^CD14^+^ cells and the expression should correlate with *CDC42* and other genes in the signature (Fig. 2G). Hence, the correlation between the MetSig and *CDC42* expression was strong in CD14^+^ cells of both RA cohorts (BiOCURA, r=0.65, p<2e-16; NeumRA, r=0.80, p<2e-16, Fig. S2D). To identify CD14^+^ cells carrying the CDC42-related MetSig, we constructed a linear regression model between the maximal and the minimal sum of the standardized expression of the signature genes and *CDC42* expression (Fig. 2H). We observed that CD14^+^ cells in the BiOCURA cohort of patients with active RA, presented two independent groups, where the MetSig had a direct correlation with the *CDC42* gene (Fig. 2H). The CD14^+^ cells within the model had higher MetSig (*MetSig*^*hi*^, n=35) compared to those outside the model (*MetSig*^*lo*^ cells, n=42), while expression of *CDC42* between the groups was comparable (Fig. S2E). To investigate the phenotype distinctions between the *MetSig*^hi^ and *MetSig*^lo^ CD14^+^ cells, we analyzed the differentially expressed genes. In addition to translation, proteasome function and OXPHOS, the genes upregulated in *MetSig*^*hi*^CD14^+^ cells were functional in *Antigen presentation* (GO:0019882, FDR=1.72e^-06^) and *Leukocyte chemotaxis* (GO:0030595, FDR=3.49e^-05^) (Fig. 2I). The antigen presentation was reflected by numerous HLA genes including the strongest known RA-risk gene *HLA-DRB1*, and β2-microglobulin (*B2M), TAP1* and invariant chain *CD74/CLIP* (Fig. 2J). Furthermore, the proteasome complex was enriched with the immunoproteasome subunits *PSMB8, PSMB10* and *PSME1*, consistent with acquisition of the antigen presenting function by the *MetSig*^*hi*^CD14^+^ cells (Fig. 2L). The *MetSig*^*hi*^CD14^+^ cells produced a broad spectrum of pro-inflammatory C-C and C-X-C chemokines including CCL2, CCL3, CCL8 and CXCL8, CXCL10 as well as the cytokines IL-1β, IL-6 and IL-10 (Fig. 2K). This abundance in chemoattractants lent support to a connection between the metabolic signature and the migratory phenotype of *CDC42*^*hi*^CD14^+^ cells and reflected the functional importance of these cells in the Rho-GTPase dependent inflammation in RA.

### CD14^+^ cells with high metabolic signature migrated into synovial tissue in RA

To investigate if *CDC42*^*hi*^CD14^+^ cells were committed to migrate into inflamed joints in RA, we turned our attention to a recent atlas of the CD11b^+^CD64^+^HLA-DR^+^ synovial tissue macrophages (STM) at the level of single-cell resolution^32^. First, we utilized the Uniform Manifold Approximation and Projection (UMAP) analysis to find the STM clusters with high RNA levels of *CDC42*. Applying the expression of *CDC42* and the MetSig genes, we identified the STM cluster which combined the high *CDC42* expression and high MetSig and, in this respect, had remarkable similarity with the blood *CDC42*^*hi*^CD14^+^ cells (Fig. 3A & Fig. S3A-B). In the next step, we compared the transcriptomics of *CDC42*^*hi*^STM cluster to the remaining cells in the given scRNA-seq and extracted the unique transcriptional characteristics of this cluster. Similar to the blood *CDC42*^*hi*^CD14^+^ cells, the *CDC42*^hi^STM cluster expressed Rho-GTPases *RAC2, RHOC* and *RHOB* and was enriched with the genes operated in *Proteasome complex* and *Antigen presentation*, and with the genes activated in *Response to IFN-γ*. Similar to the blood *CDC42*^*hi*^CD14^+^ cells, the *CDC42*^hi^STM cluster accumulated the transcriptional targets of NFE2L1 and NFRKB (Fig. 3C). Specifically, *CDC42*^*hi*^STM harbored expression of the complete set of immunoproteasome subunits *PSMB8, PSMB9, PSMB10*, and PA28 proteins *PSME1* and *PSME2* (Fig. 3B). This immunoproteasome domination was combined with high mRNA levels of the HLA-A, HLA-E and HLA-DPA genes, *WAS* protein linking CDC42 to MHC complex and the peptide transporters *TAP1, TAP2, TAPBP*, and *FCGR1A* (coding for CD64) which load the peptides into MHC complex (Fig. 3B).

**Figure 3.**
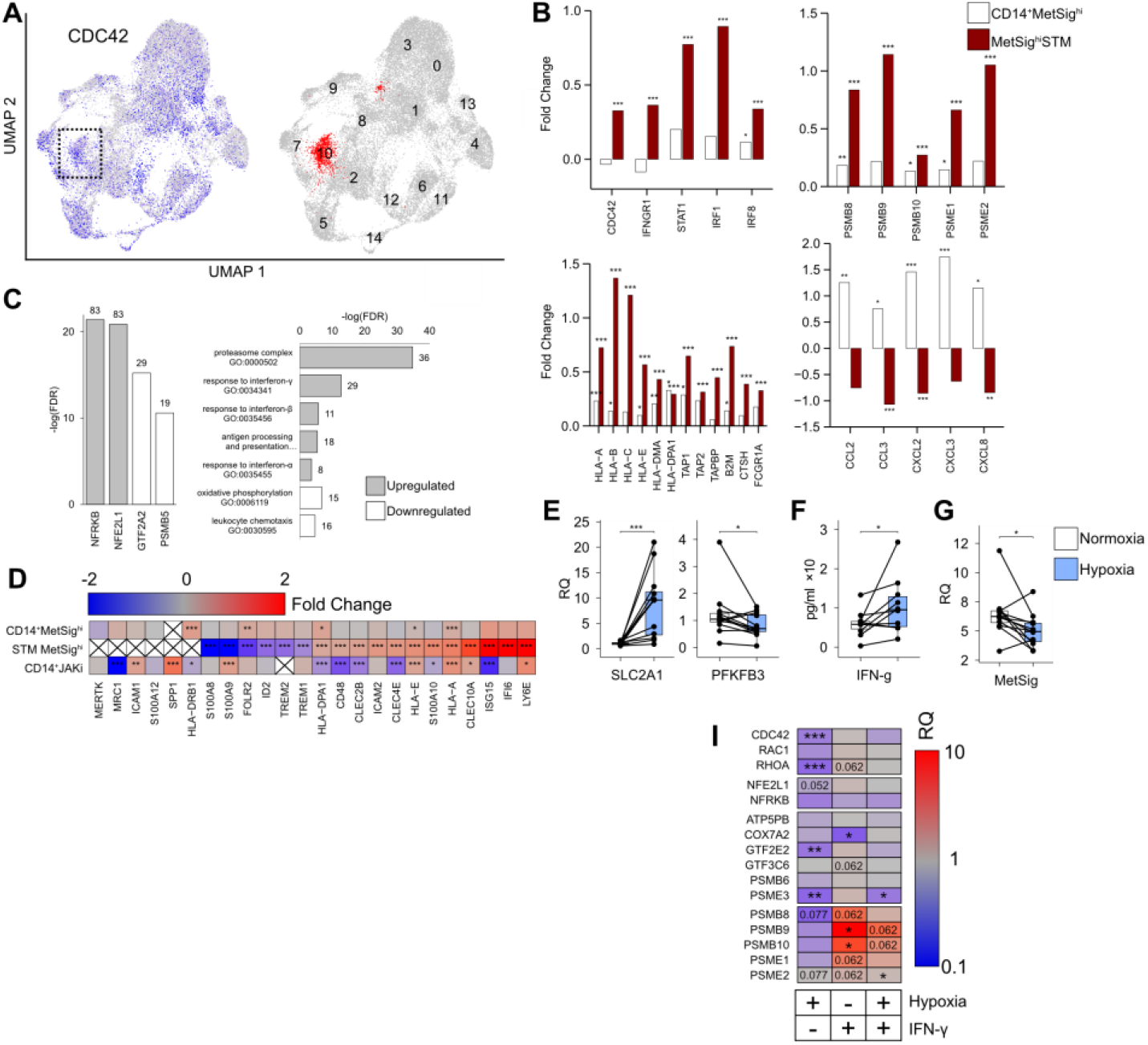
*CDC42*^*hi*^*MetSig*^*hi*^ cluster of synovial tissue macrophages (STM). **(A)** UMAP of the single-cell RNA-seq show distribution of CD11b^+^CD64^+^HLA-DR^+^ cells with high expression of *CDC42* (blue). Cluster with the highest metabolic signature (MetSig) is marked red. **(B)** Bar plot of the gene expression difference in fold change (FC) between MetSig^hi^ and MetSig^lo^ CD14^+^ cells of BiOCURA cohort (open bars) and between MetSig^hi^STM and other STM clusters (red bars). Genes involved in immunoproteasome, antigen presentation, chemokines, and response to interferon are shown separately. **(C)** Bar plots of the false discovery rate (FDR) for the GO biological processes and transcription factor (TF) targets enriched (grey bars) and downregulated (open bars) in MetSig^hi^STM cluster. **(D)** Heatmap of the gene expression difference in fold change between MetSig^hi^ and MetSig^lo^ CD14^+^ cells of BiOCURA cohort, MetSig^hi^STM and other STM clusters, and between CD14^+^ cells of JAKi-treated patients compare to other treatments in the NeumRA cohort. P-values are calculated by DESeq2 test for bulk RNA-seq and Wilcoxon rank sum test for scRNA. (*) indicates p-value<0.05. (**) indicates p-value<0.01. (***) indicates p-value<0.001. **(E)** Box plot of *SLC2A1* and *PFKFB3* mRNA in CD14^+^ cells (n=12) cultured in normoxic and hypoxic (1% O_2_) conditions for 48h. Relative quantity (RQ) was calculated in relation to *ACTB* gene. **(F)** Box plot of IFN-γ protein levels (by ELISA) in supernatants of CD14^+^ cells (n=10) cultured in normoxic and hypoxic conditions. **(G)** Box plot of the MetSig genes expression in CD14^+^ cell (n=12) cultured in normoxic and hypoxic conditions. Relative quantity was calculated in relation to ACTB gene. **(I)** Heatmap of the gene expression difference in RQ between CD14^+^ cells (n=12) cultured in hypoxic conditions with IFN-γ (0 and 50 ng/ml). Expression is normalized to CD14^+^ cells cultured in normoxic conditions without IFN-γ. P-values are calculated by Wilcoxon signed rank test. (*) indicates p-value<0.05. (**) indicates p-value<0.01. (***) indicates p-value<0.001.

The top upregulated genes in the *CDC42*^*hi*^STM cluster were the IFN-γ sensitive genes *ISG15* and *IFI6*, and *Ly6E* (Fig. 3D), which revealed that *CDC42*^*hi*^STM represented IFN-γ activated cells with a consecutive expression of the *IFNGR1, STAT1, IRF1* and *IRF8* genes mediating IFN-γ signal and numerous other IFN-γ target genes including *CDC42* (Fig. S3F). The combination IFN-sensitive genes with high expression of C-type lectin receptors *CLEC10A, CLEC2B* and *CLEC4E* and low expression of the *FOLR2, ID2* and *TREM1* genes typical for the resident STM (Fig. 3D) presented a highly inflammatory subset of STM operative in autoimmune inflammation and immune cell communication^32, 42-44^. Analyzing the downregulated genes, we found enrichment for the GTF2A2 and PSMB5 transcription targets as well as for the biological processes of *Translation*, and *Leukocyte chemotaxis* (Fig. 3C). CDC42^hi^STM had suppression of CCL2, CCL3, CXCL2, CXCL3 and CXCL8 cytokines, which was in opposite to the blood CDC42^hi^CD14^+^ cells marked by the high MetSig. Together, the comparison of blood CD14^+^ cells and STM marked by the MetSig argued in favor of functional transition of the CDC42^hi^ cells from the migration-supporting chemokine production to the antigen presentation commitment after reaching synovium.

### Effect of oxygen deprivation and IFN-γ exposure on the metabolic signature of CD14^+^ cells

The inflamed RA synovium is hypoxic^9,10^. Consistent with oxygen deprivation, we found the downregulation of the mitochondrial gene expression in the *CDC42*^*hi*^ STM and a decrease in the OXPHOS (Fig. S3C). Thus, we asked if the oxygen deprivation was responsible for the observed transition from the constitutive to immunoproteasome. To investigate this, we cultured CD14^+^ cells under normoxic (21% O_2_) and hypoxic (1% O_2_) conditions. Hypoxia induced an expected increase of *SLC2A1* mRNA coding for the glucose transporter GLUT1; down-regulated the *PFKFB3* gene (Fig. 3E) characteristic for anaerobic glycolysis and caused an increase in IFN-γ production by the cultured CD14^+^ cells (Fig. 3F). Consistent with a decrease in OXPHOS, the expression of *NFE2L1, PSMB5* and *GTF2A2* TFs was suppressed in hypoxic CD14^+^ cells (Fig. 3H, Fig. S3E) and was followed by a significant suppression of the MetSig (Fig. 3G), and *CDC42* and *RhoA* GTPases (Fig. 3I). However, the short-term hypoxia was not sufficient to significantly change the expression of immunoproteasome subunits in CD14^+^ cells (Fig. 3H). In contrast, stimulation of CD14^+^ cells with IFN-γ in normoxic conditions, significantly increased expression of the immunoproteasome genes *PSMB9* and *PSMB10*, while *PSMB8, PSME1* and *PSME2* tended to decrease (Fig. 3H). Similar effect of IFN-γ stimulation, but less pronounced, was seen in CD14^+^ cells cultured under the hypoxic conditions.

Together, these results presented the experimental evidence that oxygen deprivation created the IFN-γ rich environment and suppressed the *CDC42*-related MetSig in CD14^+^ cells. The IFN-γ rich conditions were required to induce immunoproteasome expression. In the view of similarity between *CDC42*^hi^CD14^+^ and *CDC42*^hi^STM, these findings suggest that *CDC42*^hi^ cells bearing the MetSig were recent invaders into the synovia, not completely adapted to hypoxia.

### *CDC42*-related metabolic signature of CD14^+^ cells was associated with RA disease severity and was modulated by treatment

To investigate how phenotype of the *CDC42*^*hi*^CD14^+^ cells was applied to the activity of RA disease, we constructed a linear regression model between expression of the MetSig and the disease activity score (DAS28) as a dependent parameter (Fig. S4A). In total, 48.2% of the patients (35 of 77, BiOCURA cohort; 32 of 59, NeumRA cohort) showed a strong correlation between the MetSig in CD14^+^ cells and DAS28 (r= 0.77, p=4.6e^-15^. Fig. 4A). In agreement with nature of the cohorts, *CDC42*^hi^CD14^+^ cells of the BiOCURA patients with active disease had significantly higher DAS28 (Fig. 4A) and metabolic signature compared to the NeumRA cohort of treated patients. Analyzing the DEG in *CDC42*^*hi*^CD14^+^ cells that contributed to the RA disease activity, we identified 169 genes that correlated to DAS28 (|Spearman ρ|>0.5, 130 directly and 39 inversely) and confirmed their engagement in the processes of *Monocyte chemotaxis* and *Proteasome complex* (Fig. S4B). Among those, 97 (57.4%) genes were the transcriptional targets of NFE2L1, HOXA2 and NFRKB (Fig. S4C), which added an important link between those TFs and perpetuation of the RA disease activity.

**Figure 4.**
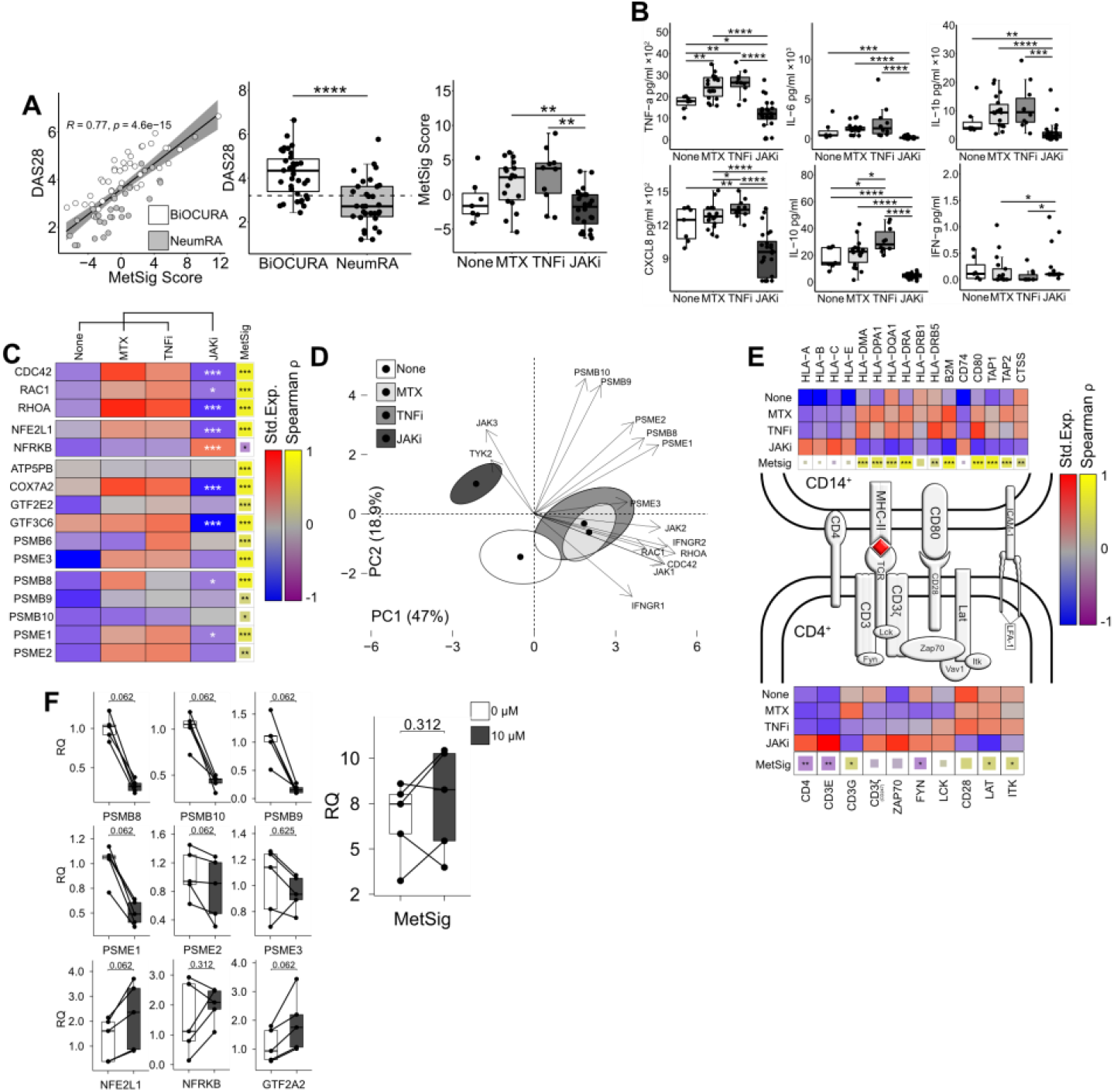
Treatment with JAK-inhibitors suppress immunoproteasome and antigen presentation in CD14^+^ cells. **(A)** Dot plot of correlation between the metabolic signature (MetSig) of CD14^+^ cells and disease activity score (DAS28) in BiOCURA and NeumRA cohorts. Box plot of DAS28 in BiOCURA and NeumRA cohorts. Box plot of MetSig in CD14^+^ cells in NeumRA cohort with different treatment. Methotrexate (MTX, n=18); TNF-a inhibitors (TNFi, n=10); JAK inhibitors (JAKi, n=24), none (n=7). **(B)** Box plots of cytokine protein production (by ELISA) by CD14^+^ cells in NeumRA cohort with different treatment and activated with LPS (5 μg/ml) for 48 h. P-values are calculated with the Mann-Whitney test. (*) indicates p-value<0.05. (**) indicates p-value<0.01. (***) indicates p-value<0.001. **(C)** Heatmap of the mean gene expression after z-transformation in CD14^+^ cells of patients with different treatment. P-values are calculated between JAKi and other treatments. Correlation between expression of individual gene and the metabolic signature is calculated by Spearman. (*) indicates p-value<0.05. (**) indicates p-value<0.01. (***) indicates p-value<0.001. **(D)** Principal component (PC) analysis of the expression of immunoproteasome genes, IFN-γ and Rho-GTPases in treatment groups. **(E)** Immunological synapse. Heatmap of the z-normalized median expression of genes active during antigen presentation in CD14^+^ cells and CD4^+^T cells. Correlation of the genes to the metabolic signature obtained by Spearman method. **(F)** Box plot of RNA levels for the immunoproteasome subunits, transcription factors and metabolic signature in CD14^+^ cells treated with tofacitinib (0 and 10 μM). P-values are calculated by paired Mann-Whitney test. **(G)** Bar plot of the expression difference in fold change (FC) of transcription factors active in CD14^+^ cell reprograming. Comparison between MetSig^hi^ and MetSig^lo^ CD14^+^ cells of BiOCURA cohort (white bars), MetSig^hi^STM and other STM clusters (red bars), and between CD14^+^ cells of JAKi-treated patients compare to other treatments in the NeumRA cohort (black bars). P-values are calculated by DESeq2 test. (*) indicates p-value<0.05. (**) indicates p-value<0.01. (***) indicates p-value<0.001.

Using the NeumRA cohort, we analyzed how RA treatment affected the CDC42-related MetSig. Overall, the CD14^+^ cells of patients treated with JAK-inhibitors (JAKi, n=24) had significantly lower the MetSig compared to CD14^+^ cells of patients treated with TNF-inhibitors (TNFi, n=10) and methotrexate (n=18), or not treated with anti-rheumatic drugs (n=7) (Fig. 4A). The MetSig in the treatment groups was mirrored by the expression of *CDC42, RAC1* and *RHOA*, transcription factor *NFE2L1* and the immunoproteasome genes in RNA-seq (Fig. 4C), and the release of IL-6, TNF-α, IL-1β, and CXCL8 by cultured CD14^+^ cells (Fig. 4B). Principal component analysis (PCA) visualized divergence between CD14^+^ cells of the treatment groups regarding the Rho-GTPase expression, immunoproteasome complex, and response to IFN-γ (Fig. 4D). The first two PCs accounted for 65.9% of the total variability. PC1 (47% of variability) was mainly explained by the Rho-GTPases and IFN response genes represented by the top contributors *CDC42, RHOA IFNGR2*. PC2 (18.9 % of variability) involved the immunoproteasome genes with the top contributors *PSMB9, PSMB10, PSME2*. To combine these findings with the results obtained in active RA of the BiOCURA cohort, we investigated the genes involved in the antigen processing and presentation. We found that CD14^+^ cells of the patients treated with JAKi had low MHC-II genes *HLA-DRB1* and *HLA-DPA1* and upregulated MHC-I genes *HLA-A* and *HLA-E* compared to those treated with methotrexate monotherapy or in combination with TNFi. In addition, the JAKi-treatment suppressed the peptide transporters *TAP1, TAP2* loading on MHCI and the cathepsin S protease gene *CTSS* essential for loading on MHC-II and upregulated the invariant chain gene *CD74*. The genes suppressed in CD14^+^ cells of the JAKi-treated patients correlated with the MetSig (Fig. 4E), which further confirmed a consistent effect of JAKi on the features related to the MetSig and lent a believe that treatment with JAKi led to a restoration of healthy synovial homeostasis. Together with the reduction in Rho-GTPases and the MetSig, the CD14^+^ cells of the JAKi-treated patients had a significantly low expression of IFN-γ -dependent MHC co-receptors *CD48, MRC1* and, also *CLEC2B, CLEC4E* and *ISG15*, while the expression of *ICAM1*, alarmin *S100A9, CLEC10A* and osteopontin gene *SPP1* remained highly expressed (Fig. 3D).

Antigen presentation through MHC class II mediates interaction between CD14^+^ and CD4^+^ T-helper cells. Hence, we examined the genes active in TCR complex in CD4^+^ cells which have been isolated simultaneously with CD14^+^ cells (Fig. 4E). The analysis revealed that JAKi-treatment resulted in upregulation of the integral components of the TCR complex *CD4*, CD3ζ (encoded by *CD247*), *ZAP70, LCK, CD3E, FYN, VAV1* and the LFA-1 subunit *ITGB2*, required for TCR activation, while the co-stimulation genes *CD28, LAT* and *ITK* were significantly down-regulated. Accordingly, *CD4, CD3E* and *FYN* were inversely correlated to the metabolic signature of CD14^+^ cells (Fig. 4E). These findings demonstrated that treatment with JAKi caused a pronounced disbalance in expression of the genes mediating interaction between CD14^+^ and CD4^+^ cells, which affected function of the immunologic synapse. This could explain the low cytokine production in CD14^+^ cells of the patients treated with JAKi (Fig. 4E), dependent on those cell interaction^45,46^.

To explore if JAKi have a direct effect on the metabolic signature and immunoproteasome production of CD14^+^ cells, we cultured freshly isolated CD14^+^ cells in presence of JAKi tofacitinib and measured expression of the immunoproteasome specific genes and the MetSig genes. We found that the CD14^+^ cells cultured with JAKi upregulated expression of the TFs *NFE2L1*, and *GTF2A2* that controlled the MetSig and down-regulated significantly expression of the immunoproteasome subunits *PSMB8, PSMB9, PSMB10, PSME1*, and *PSME2* (Fig. 4F). The MetSig of CD14^+^ cells cultured with JAKi had no significant expression change. This indicated that JAKi-inhibitors had a direct NFE2L1-dependent effect on the transcriptional regulation of immunoproteasome in CDC42^hi^CD14^+^ cells. On the one hand, the observed effect was independent of CD4^+^ cells, on the other hand, it could be a prerequisite for optimal interaction between these cells within the immunologic synapsis.

## Discussion

This study shows that circulating *CDC42*^*hi*^CD14^+^ cells are characterized by a specific metabolic signature, which combines the processes of activated oxidative phosphorylation, and proteasome-dependent protein remodeling in those cells. In blood CD14^+^ cells of RA patients, the metabolic signature included expression of the *ATP5BP, COX7A2, PSMB6, PSME3, GTF3C6*, and *GTF2E2* genes, which individually correlated to *CDC42*, and together, identified the patients where clinical RA disease activity was dependent on the CDC42-related metabolic signature of CD14^+^ cells. We observed that the high metabolic signature was functionally translated into antigen presentation. Indeed, these cells expressed multiple MHC molecules, their co-receptor b2-microglobulin and antigen-associated invariant chain CD74/CLIP, and the antigen loading proteins TAP1 and TAP2. The *CDC42*^hi^CD14^+^ cells marked by the high metabolic signature produced pro-inflammatory cytokines and a broad spectrum of chemokines, presenting a pattern of signal molecules typical to recruit immune cells to the inflamed joints in RA^2,47^. In addition to being the strongest RA risk factor, MHC molecules distinguished *CDC42*^hi^CD14^+^ cells with efficient antigen-presenting ability that accumulate the signals and mediate them further coordinating activity of the innate and adaptive immunity at the site of inflammation.

This study identified the *CDC42*^hi^CD14^+^ cells to be ancestors of the tissue infiltrating *CDC42*^hi^STM, which were phenotypically united by the metabolic signature, expression of MHC receptors and the proteasome-dependent protein remodeling. Consistent with the recent extensive analysis of the HLA-DR^+^ STM^48^, the *CDC42*^hi^STM were characterized by a suppression of the tissue-resident macrophage marks *FLOR2, ID2, TREM2*, while the IFN-sensitive genes *ISG15, CLEC10A* and *CD47* were highly expressed and revealed an invasive nature of the *CDC42*^hi^STM ready for maintaining active synovial inflammation. In the environment of synovial tissue, these *CDC42*^hi^STM gained a unique expression of the complete set of immunoproteasome genes, and the antigen presenting function of MHC was enhanced by expression of a high affinity Fc-g receptor *FCGR1A* and C-type lectin receptors to coordinate Rho-GTPase dependent antigen uptake^49,50^ attracting the adaptive immune responses into the RA synovium. A link between high C-lectin expression in CD14^+^ cells and autoimmunity has been shown in the clinical setting and experimentally for RA^51^ and multiple sclerosis^42^.

Proteasome has emerged as a crucial regulator of macrophage plasticity that maintained proteostasis in challenged cells and tissues^52-54^. Conversion to immunoproteasome helps cell to handle the excess of damages proteins by significantly increasing the substrate turnover capacity^55^. In patients with active RA disease, we observed a general increase in expression of proteasome proteins in blood *CDC42*^hi^CD14^+^, which in *CDC42*^hi^STM, was fortified further by a significant enrichment for immunoproteasome without reduction in constitutive proteasome. Turnover of Rho-GTPases in the activated cells is processed through proteasome^56^. Thus, constitutive activation of Rho-GTPases in RA *CDC42*^hi^CD14^+^ cells and STM, could have been triggered expression of the immunoproteasome proteins in similarities to that described during infection^57,58^.

Synovial hypoxia and mitochondrial dysfunction that maintain inflammation in RA^59-61^, has been shown to induces a transition from constitutive to immunoproteasome^62^. This was followed by the upregulation of MHCII antigens and acquisition of an immunogenic cell phenotype. We observed that the experimental hypoxia alone was not sufficient to reproduce the phenotype of *CDC42*^hi^STM by upregulating the immunoproteasome complex despite that it led to downregulation of the OXPHOS-based metabolic signature of CD14^+^ cells, upregulation of glucose consumption, and increased IFN-γ production. These observations led us to suggest that the cluster of *CDC42*^hi^STM was rather affected by the IFN-rich environment than by tissue hypoxia. which also caused a conversion from canonical to immunoproteasome production and increased expression of the numerous IFN-sensitive genes. Thus, entering synovia *CDC42*^hi^CD14^+^ cells were transformed into mediators of immunological information with a strong potential to shape biological processes in RA synovium, as we have reported in the GLC mice^18,23^.

In *CDC42*^*hi*^CD14^+^ cells of RA patients the OXPHOS was controlled by NFE2L1, and NFRKB TFs and supplied these cells with the energy required for the chemokine guided migration. NFE2L1 and its paralog NFE2L3 maintain the basal proteasome activity, while simultaneous deletion of those proteins impairs proteasome function in cancer cells^39,63^. Acting through down-regulation of NFE2L1, autoimmune inflammation has been shown to trigger the subunit displacement in S26 proteasome complex forming the immunoproteasome^64^. Indeed, we found that experimental hypoxia suppressed mRNA levels of *NFE2L1* in CD14^+^ cells having no significant effect on the metabolic signature. In contrast, CD14^+^ cells cultured with JAKi upregulated *NFE2L1*, which could have caused the observed decrease of immunoproteasome subunits. Despite that we observed no significant change in *NFE2L1* and *NFRKB* in *CDC42*^hi^STM and after IFN-γ stimulation, this presented a plausible checkpoint that controlled proteasome enrichment in *CDC42*^hi^CD14^+^ cells and enhance it under the conditions of chronic IFN-γ stimulation after the synovial tissue influx.

The results of our study showed that IFN-rich environment is a critical parameter for the phenotype of *CDC42*^hi^STM after they have left circulation. Consistently, treatment of RA patients with JAKi abrogated IFN signaling and suppressed the metabolic signature, which led to dramatic results changing the phenotype of CD14^+^ cells. These CD14^+^ cells down-regulated the immunoproteasome genes, Rho-GTPases, and the MHCII genes, followed by disintegration of contact with CD4^+^ cells and suppressed production of proinflammatory cytokines, potentially limiting synovial infiltration. The effects achieved by JAKi were similar to those reported for the proteasome inhibitors in patients with autoimmune diseases and in animal models of autoimmunity^65^. Success of JAKi in treatment of the autoinflammatory syndrome caused the gain-of-function mutation in PSMB8 and in interferonopathies further stress the pathogenetic closeness of these conditions^66-68^. Analogously, JAKi have been shown efficient in controlling disease progress in multiple myeloma, where proteasome inhibitors are currently used as the first line treatment^69^.

Taken together, this study demonstrates that *CDC42*^hi^CD14^+^ cells are precursors of a STM cell subset with strong antigen-presenting capacity that shapes and directs the adaptive immune responses in autoimmune inflammation. The study exhibits the important role of immunoproteasome in homeostasis of those pathogenic macrophages. The CDC42-related MetSig of CD14^+^ cells identified a substantial group of RA patients in whom these molecular processes were associated and maintained the disease activity. This group of RA patients was sensitive to treatment with JAKi and responded the treatment with downregulation of Rho-GTPases and immunoproteasome subunits, which resulted in a loss of interaction in the immunological synapse, and in a decrease in tissue-infiltrating macrophage population. The reversible and JAKi sensitive nature of the *CDC42*^hi^CD14^+^ cells predicts that RA patients carrying the *CDC42*-related MetSig may favor of early use of this targeted intervention, which justifies its evaluation prior to treatment choice.

## Data Availability

All data produced in the present study are available upon reasonable request to the authors

## Conflict of Interest

The authors declare that the research was conducted in the absence of any commercial or financial relationships that could be construed as a potential conflict of interest.

## Author Contributions

**E.M.B. & M.I.B:** conceived the project and experiments, **S.T.S**., **R.P**., **M.I.B**. provided samples, equipment, and reagents. **E.M.B**., **K.M.E.A. & M.C.E** performed experiments and analyzed the data.

**E.M.B. & M.I.B** wrote the manuscript. All authors contributed to the article and approved the submitted version.

## Fundings

This work has been funded by grants from the Swedish Research Council (MB, 2017-03025 and 2017-00359), the Swedish Association against Rheumatism (MB, R-566961, R-751351 and R-860371), the King Gustaf V:s 80-year Foundation (MB, FAI-2018-0519 and FAI-2020-0653), the Regional agreement on medical training and clinical research between the Western Götaland county council and the University of Gothenburg (MB, ALFGBG-717681, ALFGBG-965623; RP, ALFGBG-965012, ALFGBG-926621), the University of Gothenburg. The authors declare that the funding sources have no role in study design; in the collection, analysis, and interpretation of data; in the writing of the report; and in the decision to submit the paper for publication.

## Acknowledgments

We would like to thank the research nurses Anneli Lund and Marie-Louise Andersson at the Rheumatology Clinic, Sahlgrenska University Hospital, Gothenburg, for their help with blood sampling. We also thank all RA patients, who participated in this study. We appreciate support of Aridaman Pandit and Weiyang Tao at the Center for Translational Immunology, University Medical Center Utrecht, The Netherlands, for sharing clinical data of the BiOCURA cohort. We are thankful to Prof. Marcela Pekna at the Department of Clinical Neuroscience, University of Gothenburg, for assistance with the hypoxia experiments.

## Supplementary Material

Supplemental Table S1 and S2 and Supplemental Figures 1-4 are given in the supplementary material document.

## Data Availability Statement

Bulk RNA-seq data have been deposited at GEO and are currently publicly available. Accession numbers are GSE201670 and GSE201669. This paper analyzes existing, publicly available data. These accession numbers for the datasets are listed in the material and methods section. This paper does not report original code. Any additional information required to reanalyze the data reported in this paper is available from the lead contact upon request.

## Notes

### Competing Interest Statement

The authors have declared no competing interest.

### Funding Statement

This work has been funded by grants from the Swedish Research Council (MB, 2017-03025 and 2017-495 00359), the Swedish Association against Rheumatism (MB, R-566961, R-751351 and R-860371), the 496 King Gustaf V:s 80-year Foundation (MB, FAI-2018-0519 and FAI-2020-0653), the Regional 497 agreement on medical training and clinical research between the Western Gotaland county council and 498 the University of Gothenburg (MB, ALFGBG-717681, ALFGBG-965623; RP, ALFGBG-965012, 499 ALFGBG-926621), the University of Gothenburg. The authors declare that the funding sources have 500 no role in study design; in the collection, analysis, and interpretation of data; in the writing of the 501 report; and in the decision to submit the paper for publication.

### Author Declarations

The study was approved by the Swedish Ethical Review Authority (659-2011) and done in accordance with the Declaration of Helsinki. The trial is registered at ClinicalTrials.gov (ID NCT03449589)

## References

1. Udalova IA, Mantovani A, Feldmann M. Macrophage heterogeneity in the context of rheumatoid arthritis. Nature Rev Rheumatol. 2016;12(8):472–85.

2. Firestein GS, McInnes IB. Immunopathogenesis of Rheumatoid Arthritis. Immunity. 2017;46(2):183–96.

3. Herenius MM, Thurlings RM, Wijbrandts CA, Bennink RJ, Dohmen SE, Voermans C, et al. Monocyte migration to the synovium in rheumatoid arthritis patients treated with adalimumab. Ann Rheum Dis. 2011;70(6):1160–2.

4. Smiljanovic B, Radzikowska A, Kuca-Warnawin E, Kurowska W, Grun JR, Stuhlmuller B, et al. Monocyte alterations in rheumatoid arthritis are dominated by preterm release from bone marrow and prominent triggering in the joint. Ann Rheum Dis. 2018;77(2):300–8.

5. Gomez EA, Colas RA, Souza PR, Hands R, Lewis MJ, Bessant C, et al. Blood pro-resolving mediators are linked with synovial pathology and are predictive of DMARD responsiveness in rheumatoid arthritis. Nat Commun. 2020;11(1):5420.

6. Jakubzick CV, Randolph GJ, Henson PM. Monocyte differentiation and antigen-presenting functions. Nat Rev Immunol. 2017;17(6):349–62.

7. Wehr P, Purvis H, Law SC, Thomas R. Dendritic cells, T cells and their interaction in rheumatoid arthritis. Clin Exp Immunol. 2019;196(1):12–27.

8. McGarry T, Hanlon MM, Marzaioli V, Cunningham CC, Krishna V, Murray K, et al. Rheumatoid arthritis CD14(+) monocytes display metabolic and inflammatory dysfunction, a phenotype that precedes clinical manifestation of disease. Clin Transl Immunology. 2021;10(1):e1237.

9. Ng CT, Biniecka M, Kennedy A, McCormick J, Fitzgerald O, Bresnihan B, et al. Synovial tissue hypoxia and inflammation in vivo. Ann Rheum Dis. 2010;69(7):1389–95.

10. Quinonez-Flores CM, Gonzalez-Chavez SA, Pacheco-Tena C. Hypoxia and its implications in rheumatoid arthritis. J Biomed Sci. 2016;23(1):62.

11. Pinet V, Vergelli M, Martin R, Bakke O, Long EO. Antigen presentation mediated by recycling of surface HLA-DR molecules. Nature. 1995;375(6532):603–6.

12. Veerappan Ganesan AP, Eisenlohr LC. The elucidation of non-classical MHC class II antigen processing through the study of viral antigens. Curr Opin Virol. 2017;22:71–6.

13. van den Eshof BL, Medfai L, Nolfi E, Wawrzyniuk M, Sijts A. The Function of Immunoproteasomes-An Immunologists’ Perspective. Cells. 2021;10(12).

14. Cascio P. PA28gamma: New Insights on an Ancient Proteasome Activator. Biomolecules. 2021;11(2).

15. Bussi C, Heunis T, Pellegrino E, Bernard EM, Bah N, Dos Santos MS, et al. Lysosomal damage drives mitochondrial proteome remodelling and reprograms macrophage immunometabolism. Nat Commun. 2022;13(1):7338.

16. Kruger E, Kloetzel PM. Immunoproteasomes at the interface of innate and adaptive immune responses: two faces of one enzyme. Curr Opin Immunol. 2012;24(1):77–83.

17. Khan OM, Akula MK, Skalen K, Karlsson C, Stahlman M, Young SG, et al. Targeting GGTase-I activates RHOA, increases macrophage reverse cholesterol transport, and reduces atherosclerosis in mice. Circulation. 2013;127(7):782–90.

18. Khan OM, Ibrahim MX, Jonsson IM, Karlsson C, Liu M, Sjogren AK, et al. Geranylgeranyltransferase type I (GGTase-I) deficiency hyperactivates macrophages and induces erosive arthritis in mice. J Clin Invest. 2011;121(2):628–39.

19. Laudanna C, Campbell JJ, Butcher EC. Role of Rho in chemoattractant-activated leukocyte adhesion through integrins. Science. 1996;271(5251):981–3.

20. Saoudi A, Kassem S, Dejean A, Gaud G. Rho-GTPases as key regulators of T lymphocyte biology. Small GTPases. 2014;5.

21. Shurin GV, Tourkova IL, Chatta GS, Schmidt G, Wei S, Djeu JY, et al. Small rho GTPases regulate antigen presentation in dendritic cells. J Immunol. 2005;174(6):3394–400.

22. Schulz AM, Stutte S, Hogl S, Luckashenak N, Dudziak D, Leroy C, et al. Cdc42-dependent actin dynamics controls maturation and secretory activity of dendritic cells. J Cell Biol. 2015;211(3):553–67.

23. Malmhall-Bah E, Andersson KME, Erlandsson MC, Akula MK, Brisslert M, Wiel C, et al. Rho-GTPase dependent leukocyte interaction generates pro-inflammatory thymic Tregs and causes arthritis. J Autoimmun. 2022;130:102843.

24. Lopez-Posadas R, Fastancz P, Martinez-Sanchez LDC, Panteleev-Ivlev J, Thonn V, Kisseleva T, et al. Inhibiting PGGT1B Disrupts Function of RHOA, Resulting in T-cell Expression of Integrin alpha4beta7 and Development of Colitis in Mice. Gastroenterology. 2019;157(5):1293–309.

25. Akula MK, Ibrahim MX, Ivarsson EG, Khan OM, Kumar IT, Erlandsson M, et al. Protein prenylation restrains innate immunity by inhibiting Rac1 effector interactions. Nat Commun. 2019;10(1):3975.

26. Manresa-Arraut A, Johansen FF, Brakebusch C, Issazadeh-Navikas S, Hasseldam H. RhoA Drives T-Cell Activation and Encephalitogenic Potential in an Animal Model of Multiple Sclerosis. Front Immunol. 2018;9:1235.

27. Ng SY, Brown L, Stevenson K, deSouza T, Aster JC, Louissaint A, et al. RhoA G17V is sufficient to induce autoimmunity and promotes T-cell lymphomagenesis in mice. Blood. 2018;132(9):935–47.

28. Du X, Zeng H, Liu S, Guy C, Dhungana Y, Neale G, et al. Mevalonate metabolism-dependent protein geranylgeranylation regulates thymocyte egress. J Exp Med. 2020;217(2).

29. Guo F, Zhang S, Tripathi P, Mattner J, Phelan J, Sproles A, et al. Distinct roles of Cdc42 in thymopoiesis and effector and memory T cell differentiation. PLoS One. 2011;6(3):e18002.

30. Aletaha D, Neogi T, Silman AJ, Funovits J, Felson DT, Bingham CO, 3rd, et al. 2010 Rheumatoid arthritis classification criteria: an American College of Rheumatology/European League Against Rheumatism collaborative initiative. Arthritis Rheum. 2010;62(9):2569–81.

31. Tao W, Concepcion AN, Vianen M, Marijnissen ACA, Lafeber F, Radstake T, et al. Multiomics and Machine Learning Accurately Predict Clinical Response to Adalimumab and Etanercept Therapy in Patients With Rheumatoid Arthritis. Arthritis Rheumatol. 2021;73(2):212–22.

32. Alivernini S, MacDonald L, Elmesmari A, Finlay S, Tolusso B, Gigante MR, et al. Distinct synovial tissue macrophage subsets regulate inflammation and remission in rheumatoid arthritis. Nat Med. 2020;26(8):1295–306.

33. Andersson KM, Turkkila M, Erlandsson MC, Bossios A, Silfversward ST, Hu D, et al. Survivin controls biogenesis of microRNA in smokers: A link to pathogenesis of rheumatoid arthritis. Biochim Biophys Acta Mol Basis Dis. 2017;1863(3):663–73.

34. Love MI, Huber W, Anders S. Moderated estimation of fold change and dispersion for RNA-seq data with DESeq2. Genome Biol. 2014;15(12):550.

35. Raudvere U, Kolberg L, Kuzmin I, Arak T, Adler P, Peterson H, et al. g:Profiler: a web server for functional enrichment analysis and conversions of gene lists (2019 update). Nucleic Acids Res. 2019;47(W1):W191–98.

36. Subramanian A, Tamayo P, Mootha VK, Mukherjee S, Ebert BL, Gillette MA, et al. Gene set enrichment analysis: A knowledge-based approach for interpreting genome-wide expression profiles. 2005;102(43):15545–50.

37. Kolmykov S, Yevshin I, Kulyashov M, Sharipov R, Kondrakhin Y, Makeev VJ, et al. GTRD: an integrated view of transcription regulation. Nucleic Acids Res. 2021;49(D1):D104–d11.

38. Hao Y, Hao S, Andersen-Nissen E, Mauck WM, Zheng S, Butler A, et al. Integrated analysis of multimodal single-cell data. Cell. 2021;184(13):3573-87.e29.

39. Kim HM, Han JW, Chan JY. Nuclear Factor Erythroid-2 Like 1 (NFE2L1): Structure, function and regulation. Gene. 2016;584(1):17–25.

40. Brotto DB, Siena ADD, de B, II, Carvalho S, Muys BR, Goedert L, et al. Contributions of HOX genes to cancer hallmarks: Enrichment pathway analysis and review. Tumour biology : the journal of the International Society for Oncodevelopmental Biology and Medicine. 2020;42(5):1010428320918050.

41. Peng Q, Wan D, Zhou R, Luo H, Wang J, Ren L, et al. The biological function of metazoan-specific subunit nuclear factor related to kappaB binding protein of INO80 complex. Int J Biol Macromol. 2022;203:176–83.

42. N’Diaye M, Brauner S, Flytzani S, Kular L, Warnecke A, Adzemovic MZ, et al. C-type lectin receptors Mcl and Mincle control development of multiple sclerosis-like neuroinflammation. J Clin Invest. 2020;130(2):838–52.

43. Welte S, Kuttruff S, Waldhauer I, Steinle A. Mutual activation of natural killer cells and monocytes mediated by NKp80-AICL interaction. Nat Immunol. 2006;7(12):1334–42.

44. Wu X, Liu Y, Jin S, Wang M, Jiao Y, Yang B, et al. Single-cell sequencing of immune cells from anticitrullinated peptide antibody positive and negative rheumatoid arthritis. Nat Commun. 2021;12(1):4977.

45. Agbanoma G, Li C, Ennis D, Palfreeman AC, Williams LM, Brennan FM. Production of TNF-alpha in macrophages activated by T cells, compared with lipopolysaccharide, uses distinct IL-10-dependent regulatory mechanism. J Immunol. 2012;188(3):1307–17.

46. Burger D, Dayer JM. The role of human T-lymphocyte-monocyte contact in inflammation and tissue destruction. Arthritis Res. 2002;4 Suppl 3(Suppl 3):S169–76.

47. Miyabe Y, Lian J, Miyabe C, Luster AD. Chemokines in rheumatic diseases: pathogenic role and therapeutic implications. Nature Rev Rheumatol. 2019;15(12):731–46.

48. Kurowska-Stolarska M, Alivernini S. Synovial tissue macrophages in joint homeostasis, rheumatoid arthritis and disease remission. Nature Rev Rheumatol. 2022;18(7):384–97.

49. Goodridge HS, Underhill DM, Touret N. Mechanisms of Fc receptor and dectin-1 activation for phagocytosis. Traffic. 2012;13(8):1062–71.

50. Dhodapkar KM, Krasovsky J, Williamson B, Dhodapkar MV. Antitumor monoclonal antibodies enhance cross-presentation ofcCellular antigens and the generation of myeloma-specific killer T cells by dendritic cells. J Exp Med. 2002;195(1):125–33.

51. Erlandsson MC, Erdogan S, Wasen C, Andersson KME, Silfversward ST, Pullerits R, et al. IGF1R signalling is a guardian of self-tolerance restricting autoantibody production. Front Immunol. 2022;13:958206.

52. Kotschi S, Jung A, Willemsen N, Ofoghi A, Proneth B, Conrad M, et al. NFE2L1-mediated proteasome function protects from ferroptosis. Mol Metab. 2022;57:101436.

53. Sha Z, Goldberg AL. Proteasome-mediated processing of Nrf1 is essential for coordinate induction of all proteasome subunits and p97. Curr Biol. 2014;24(14):1573–83.

54. Vangala JR, Radhakrishnan SK. Nrf1-mediated transcriptional regulation of the proteasome requires a functional TIP60 complex. J Biol Chem. 2019;294(6):2036–45.

55. Seifert U, Bialy LP, Ebstein F, Bech-Otschir D, Voigt A, Schroter F, et al. Immunoproteasomes preserve protein homeostasis upon interferon-induced oxidative stress. Cell. 2010;142(4):613–24.

56. Stubbs EB, Von Zee CL. Prenylation of Rho G-Proteins: a Novel Mechanism Regulating Gene Expression and Protein Stability in Human Trabecular Meshwork Cells. Mol Neurobiol. 2012;46(1):28–40.

57. Munro P, Flatau G, Doye A, Boyer L, Oregioni O, Mege JL, et al. Activation and proteasomal degradation of rho GTPases by cytotoxic necrotizing factor-1 elicit a controlled inflammatory response. J Biol Chem. 2004;279(34):35849–57.

58. Doye A, Mettouchi A, Bossis G, Clement R, Buisson-Touati C, Flatau G, et al. CNF1 exploits the ubiquitin-proteasome machinery to restrict Rho GTPase activation for bacterial host cell invasion. Cell. 2002;111(4):553–64.

59. Becker YLC, Duvvuri B, Fortin PR, Lood C, Boilard E. The role of mitochondria in rheumatic diseases. Nature Rev Rheumatol. 2022;18(11):621–40.

60. Kaur G, Sharma A, Bhatnagar A. Role of oxidative stress in pathophysiology of rheumatoid arthritis: insights into NRF2-KEAP1 signalling. Autoimmunity. 2021;54(7):385–97.

61. Clayton SA, MacDonald L, Kurowska-Stolarska M, Clark AR. Mitochondria as Key Players in the Pathogenesis and Treatment of Rheumatoid Arthritis. Front Immunol. 2021;12:673916.

62. Abu-El-Rub E, Sareen N, Yan W, Alagarsamy KN, Rafieerad A, Srivastava A, et al. Hypoxia-induced shift in the phenotype of proteasome from 26S toward immunoproteasome triggers loss of immunoprivilege of mesenchymal stem cells. Cell Death Dis. 2020;11(6):419.

63. Waku T, Nakamura N, Koji M, Watanabe H, Katoh H, Tatsumi C, et al. NRF3-POMP-20S Proteasome Assembly Axis Promotes Cancer Development via Ubiquitin-Independent Proteolysis of p53 and Retinoblastoma Protein. Mol Cell Biol. 2020;40(10).

64. Shanley KL, Hu CL, Bizzozero OA. Decreased levels of constitutive proteasomes in experimental autoimmune encephalomyelitis may be caused by a combination of subunit displacement and reduced Nfe2l1 expression. J Neurochem. 2020;152(5):585–601.

65. Verbrugge SE, Scheper RJ, Lems WF, de Gruijl TD, Jansen G. Proteasome inhibitors as experimental therapeutics of autoimmune diseases. Arthritis Res Ther. 2015;17(1):17.

66. Osterloh P, Linkemann K, Tenzer S, Rammensee HG, Radsak MP, Busch DH, et al. Proteasomes shape the repertoire of T cells participating in antigen-specific immune responses. Proc Natl Acad Sci U S A. 2006;103(13):5042–7.

67. Lin B, Goldbach-Mansky R. Pathogenic insights from genetic causes of autoinflammatory inflammasomopathies and interferonopathies. J Allergy Clin Immunol. 2022;149(3):819–32.

68. Miyamoto T, Honda Y, Izawa K, Kanazawa N, Kadowaki S, Ohnishi H, et al. Assessment of type I interferon signatures in undifferentiated inflammatory diseases: A Japanese multicenter experience. Front Immunol. 2022;13:905960.

69. Ghermezi M, Spektor TM, Berenson JR. The role of JAK inhibitors in multiple myeloma. Clin Adv Hematol Oncol. 2019;17(9):500–5.

